# Evaluation of SOFA-2 Score Performance Across Demographic Subgroups: An External Validation Study Using MIMIC-IV

**DOI:** 10.64898/2026.03.10.26348061

**Authors:** Jacob Ellen, Sicheng Hao, Catherine A. Gao, Maria Del Pilar Arias, Martin Viola, An-Kwok Ian Wong, Heather Mattie, William Parker, Cristina Haidau, João Matos, Renato Carneiro de Freitas Chaves, Leo Anthony Celi

## Abstract

The Sequential Organ Failure Assessment (SOFA)-2 score was recently validated for ICU mortality prediction across more than 3 million admissions but was not evaluated across demographic subgroups. We assessed the discrimination and calibration of the SOFA-2 score for ICU mortality across subgroups defined by age, sex, race and ethnicity, primary language, and insurance status. We conducted a retrospective cohort study of adult patients (aged 18 years or older) admitted to ICUs at Beth Israel Deaconess Medical Center between 2008 and 2022 (MIMIC-IV, version 3.1), selecting the first ICU admission per patient. First-day SOFA-2 scores (range, 0-24) were calculated using worst recorded values across 6 organ systems. Discrimination was assessed using AUROC, calibration using intercepts and slopes, and subgroup differences using bootstrap resampling. Among 64,015 ICU admissions (median age, 66 years [IQR, 54-78]; 56.1% male; 66.1% White), overall ICU mortality was 7.2% (n=4,596). Overall AUROC was acceptable at 0.77 (95% CI, 0.76-0.77). Notably, discrimination declined significantly with age: AUROC was 0.85 (95% CI, 0.83-0.87) for ages 18-44 and 0.72 (95% CI, 0.70-0.73) for ages 75 and older (difference in AUROC, −0.14; 95% CI, −0.16 to −0.11), with systematic underprediction of mortality in older patients (calibration intercept, 0.39). Discrimination was also significantly lower among non-English speakers (difference in AUROC, −0.04; 95% CI, −0.07 to −0.01) but did not differ significantly across documented racial and ethnic groups. Patients with unknown race/ethnicity (14.3% of the cohort) had nearly double the overall mortality rate and poor calibration. SOFA-2 demonstrated good overall performance for ICU mortality prediction but with clinically meaningful variation across demographic subgroups, particularly a substantial decline in discrimination with advancing age. These findings underscore the need for routine equity evaluation of clinical prediction tools before widespread implementation.

## Introduction

Organ dysfunction is a major contributor to ICU morbidity and mortality, and scoring systems have been developed to quantify its extent, stratify patients by risk, and track clinical trajectories.^1^ The Sequential Organ Failure Assessment (SOFA) score was developed in this context and has demonstrated strong performance in predicting ICU mortality.^2^

Since 1996, the SOFA score has been a cornerstone of critical care medicine, quantifying organ dysfunction to guide clinical decision-making,^3^ prognostication, and research.^1^ However, a growing body of evidence has raised concerns about its fairness. Several studies, particularly those focusing on critical care cohorts during the COVID-19 pandemic, have reported that the SOFA score’s predictive performance diverges across racial and ethnic groups,^4–6^ a finding of note given that illness severity scores have been proposed as a mechanism for mitigating inequities in settings of crisis care.^7^ Differences related to sex have also been reported, with women demonstrating lower SOFA scores that do not correspond to improved short-term outcomes.^8^ These findings suggest that clinical decisions relying solely on the SOFA score may not be equitable, warranting re-evaluation of its performance within diverse population subgroups.

In October 2025, an updated version of SOFA was published (SOFA-2), incorporating contemporary organ support treatments and revised thresholds across the same six organ systems.^9^ This new version was validated across more than 3 million ICU admissions from 1,319 ICUs in 9 countries and demonstrated robust predictive validity for ICU mortality (SOFA-2 AUROC, 0.79; 95% CI, 0.76-0.81).^9^ However, this effort did not include a systematic evaluation of model performance across demographic subgroups defined by sex, age, race/ethnicity, or other socially meaningful characteristics. Given that clinical prediction scores increasingly inform high-stakes decisions, understanding whether these tools perform equitably across patient populations is essential to assess their fairness and ensure they do not inadvertently perpetuate or exacerbate existing healthcare disparities.^6,10^

Therefore, we designed this study to evaluate the fairness of the SOFA-2 score using the Medical Information Mart for Intensive Care (MIMIC) IV database.^11^ We assessed model calibration and discrimination across subgroups defined by age, sex, race and ethnicity, primary language, and insurance status, given their known associations with differences in ICU mortality and care delivery.^4–6,8,12–14^ Fairness was operationalized as the equivalence in model discrimination and calibration across demographic subgroups.

## Methods

### Study design and data source

We conducted a retrospective cohort study using the Medical Information Mart for Intensive Care database (MIMIC-IV, version 3.1).^11^ This database contains deidentified data for patients admitted to ICUs at Beth Israel Deaconess Medical Center in Boston, Massachusetts, between 2008 and 2022. Since MIMIC IV was not used in the development and validation of the SOFA-2 score^9^ this study represents an external validation.

### Study population

We included hospitalized adult patients (age ≥18 years) with at least one ICU admission. We selected the first hospitalization with an ICU visit for each patient, and within that hospitalization, only the first ICU admission was analyzed. Patients were excluded if physiologic values fell outside clinically plausible ranges informed by the original SOFA-2 validation study (eTable 1)^9^ or if ICU length-of-stay was less than 6 hours.

### SOFA-2 Score calculation

SOFA-2 scores were calculated according to the methods described in the original validation study.^9^ We used the worst recorded value during the first 24 hours of ICU admission for each of the 6 organ systems: neurological (Glasgow Coma Scale), cardiovascular (mean arterial pressure and vasopressor administration), respiratory (PaO₂/FiO₂ ratio and mechanical ventilation status), hepatic (bilirubin), renal (creatinine and urine output), and coagulation (platelet count). Organ-specific subscores ranged from 0 to 4, and total SOFA-2 scores ranged from 0 to 24, with higher scores indicating more severe organ dysfunction. Missing SOFA-2 components were assigned a score of 0 (normal organ function), consistent with the original SOFA-2 validation.^9^ We analyzed only first-day SOFA-2 scores, as our primary aim was to assess potential bias during initial triage and risk stratification.

Key SOFA-2 modifications relative to the original SOFA score, as specified by Ranzani et al.,^9^ included revised PaO_2_/FiO_2_ thresholds, with SpO_2_/FiO_2_ used as a fallback when SpO_2_ was less than 98%. The respiratory component also assigned maximum scores for advanced ventilatory support modalities, including invasive mechanical ventilation, NIV, CPAP, BiPAP, HFNC, and ECMO. The cardiovascular component assigned maximum scores for mechanical circulatory support devices and applied graduated mean arterial pressure thresholds when vasopressors were not administered. Vasopressor infusions lasting at least 60 minutes were included in scoring, and additional agents considered in our implementation included milrinone, vasopressin, and phenylephrine. The neurologic component assigned a minimum score of 1 for pharmacologic treatment of delirium; qualifying medications included dexmedetomidine, haloperidol, quetiapine, ziprasidone, and olanzapine. When the total Glasgow Coma Scale (GCS) score was unavailable, the motor component was used as a fallback. The renal component assigned maximum scores for renal replacement therapy or biochemical criteria indicating its initiation, and platelet and bilirubin thresholds were also revised.

The SOFA-2 specification did not explicitly define qualifying delirium agents or vasopressors, and thus the medications above were selected following deliberation with critical care physicians. Some features specified in SOFA-2 could not be reliably abstracted from the electronic health record (see Limitations)

### Demographic subgroups

Age was categorized into four groups: 18–44, 45–64, 65–74, and ≥75 years. Sex was classified as male or female based on recorded values in MIMIC-IV. Race/ethnicity was grouped as White, Black, Hispanic, Asian, Other, or Unknown if labeled as unknown, unable to obtain, or patient declined. Patients identifying as Native Hawaiian/Pacific Islander, American Indian/Alaska Native, or multiple races were combined into Other due to small sample sizes. Primary language was categorized as English, Non-English, or Unknown. Insurance was categorized as private, Medicare, Medicaid, or other.

### Outcome

The primary study outcome was ICU mortality, defined as death during the index ICU admission or within 6 hours of ICU discharge. ICU mortality was chosen to enable direct comparison with previous approaches.^1,9^

### Statistical analysis

Continuous variables were reported as median (IQR) or mean (SD) according to their distribution and categorical variables as counts (percentages). We evaluated SOFA-2 performance in the overall cohort and within subgroups using measures of discrimination and calibration. Discrimination was assessed using the area under the receiver operating characteristic curve (AUROC) with 95% confidence intervals (CIs). For interpretive context, AUROC values between 0.70 and 0.80 were considered acceptable discrimination and values exceeding 0.80 were considered good, consistent with commonly used benchmarks.^15^ Calibration was assessed using intercepts and slopes, where values of 0 and 1, respectively, indicate perfect calibration. Calibration plots were generated using predicted risk from a logistic regression model fit in the overall cohort with ICU mortality as the outcome and first-day SOFA-2 score as the sole predictor. Overall calibration metrics are expected to approximate ideal values because the calibration model was fit to the same cohort; the primary analytic interest lies in subgroup-level departures from these benchmarks.

Subgroup differences were compared using nonparametric bootstrap resampling (1,000 iterations) to obtain 95% CIs for differences in AUROC; differences were considered statistically significant if the 95% CI excluded zero. We considered ΔAUROC values exceeding 0.05 to represent potentially clinically meaningful differences.^16^ We also described observed ICU mortality rates across individual SOFA-2 scores (0–14, ≥15 combined) stratified by demographic subgroups.

All analyses used R version 4.5.0 following TRIPOD+AI guidelines (Supplemental).^17^ The complete analytic pipeline is publicly available at: https://github.com/SichengH/SOFA2_bias.

### Ethics and approval

This study used deidentified, publicly available data from the MIMIC-IV database, hosted on PhysioNet.^18^ Access was obtained after completion of required human-subjects training and user credentialing.

## Results

Of the 94,458 ICU admissions in MIMIC-IV, 65,366 met inclusion criteria. After further excluding patients with ICU stays <6 hours and implausible physiologic values (eTable 1), 64,015 ICU stays constituted the final cohort. As illustrated in Figure 1, we monitored demographic and clinical shifts throughout the exclusion process to elucidate potential sources of selection bias. Missing data proportions by SOFA-2 components are provided in eTable 2, with the hepatic component demonstrating the highest rate of missingness (58%) and the cardiovascular component the lowest (0%).

**Figure 1:**
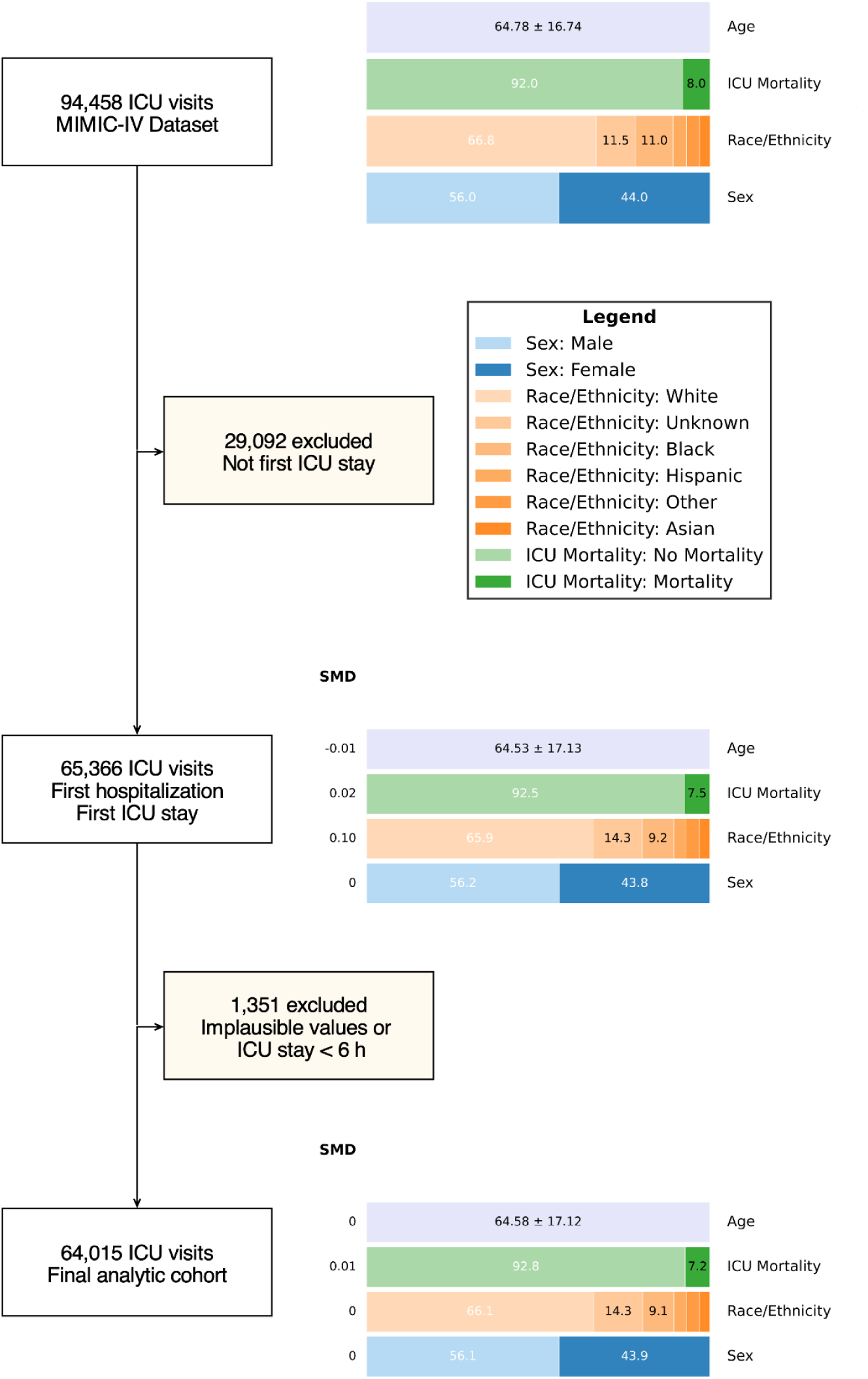
Flow diagram demonstrating the selection of a final analytic cohort. Demographic and clinical characteristics are shown at each exclusion step along with standardized mean differences (SMDs), where SMD is defined as the difference in means or proportions divided by a pooled standard deviation; values ≤ 0.10 generally indicate minimal imbalance.^31^ Age is expressed in years and reported as mean (SD); categorical variables are presented as percentages.

### Cohort characteristics

The characteristics of the study population are described in Table 1. Mean age was 64.6 years (SD 17.1), with 54.5% of patients older than 64 years. The cohort was predominantly male (56.1%), White (66.1%), and English-speaking (90.4%), with Medicare as the most common insurance type (52.8%). The median first-day SOFA-2 score was 4 (IQR, 2–6), and overall ICU mortality was 7.2% (n=4,596). The SOFA-2 score distribution is shown in eFigure 1.

**Table 1.** Baseline characteristics of the study population.

| **Characteristic** | **MIMIC-IV Patients** | **Median SOFA-2 (IQR)** | **Median LOS, days (IQR)** | **ICU Mortality, n (%)** |
| --- | --- | --- | --- | --- |
| **Overall** | 64,015 | 4 (2–6) | 1.9 (1.1–3.7) | 4,596 (7.2%) |
| **Age categories (years)** |  |  |  |  |
| 18-44 | 8428 (13.2%) | 3 (1–5) | 1.7 (1.0–3.5) | 346 (4.1%) |
| 45-64 | 20726 (32.4%) | 4 (2–6) | 1.9 (1.1–3.7) | 1,198 (5.8%) |
| 65-74 | 14735 (23.0%) | 4 (3–7) | 2.0 (1.2–3.8) | 1,019 (6.9%) |
| ≥75 | 20126 (31.4%) | 5 (3–7) | 2.0 (1.1–3.7) | 2,033 (10.1%) |
| **Sex** |  |  |  |  |
| Male | 35918 (56.1%) | 4 (2–7) | 1.9 (1.1–3.7) | 2,523 (7.0%) |
| Female | 28097 (43.9%) | 4 (2–6) | 1.9 (1.1–3.7) | 2,073 (7.4%) |
| **Race/Ethnicity** |  |  |  |  |
| White | 42338 (66.1%) | 4 (2–6) | 1.9 (1.1–3.5) | 2,544 (6.0%) |
| Black | 5833 (9.1%) | 4 (2–6) | 1.9 (1.0–3.6) | 349 (6.0%) |
| Hispanic | 2285 (3.6%) | 4 (2–6) | 1.9 (1.1–3.7) | 124 (5.4%) |
| Asian | 1925 (3.0%) | 4 (2–6) | 1.9 (1.1–3.4) | 133 (6.9%) |
| Other | 2471 (3.9%) | 4 (2–6) | 1.9 (1.1–3.9) | 153 (6.2%) |
| Unknown | 9163 (14.3%) | 5 (3–7) | 2.2 (1.2–4.9) | 1,293 (14.1%) |
| **Primary Language** |  |  |  |  |
| English | 57879 (90.4%) | 4 (2–6) | 1.9 (1.1–3.7) | 4,102 (7.1%) |
| Non-English | 5789 (9.0%) | 4 (2–6) | 2.0 (1.1–4.0) | 414 (7.2%) |
| Unknown | 347 (0.5%) | 5 (4–8) | 2.1 (1.1–5.1) | 80 (23.1%) |
| **Insurance Status** |  |  |  |  |
| Medicare | 33789 (52.8%) | 4 (3–7) | 2.0 (1.1–3.8) | 2,859 (8.5%) |
| Private | 18073 (28.2%) | 4 (2–6) | 1.8 (1.1–3.5) | 856 (4.7%) |
| Medicaid | 9149 (14.3%) | 4 (2–6) | 1.9 (1.1–3.9) | 559 (6.1%) |
| Other | 3004 (4.7%) | 4 (2–6) | 1.8 (1.0–3.5) | 322 (10.7%) |

Values are n (%) unless otherwise specified.

### Overall SOFA-2 performance

The SOFA-2 score demonstrated acceptable discrimination for ICU mortality (AUROC 0.77; 95% CI: 0.76–0.77) and calibration showing an intercept of 0.00 (95% CI: −0.03 to 0.03) and slope of 1.00 (95% CI: 0.97–1.03), consistent with the calibration model having been fit to the overall cohort. Discrimination and calibration metrics across demographic subgroups are presented in Table 2. Calibration curves are shown in eFigures 2–6, with observed mortality rates by SOFA-2 score stratified by demographic subgroups shown in Figure 2 and eFigures 7–9. Organ-specific subscores by subgroup are provided in eTable 3.

**Figure 2:**
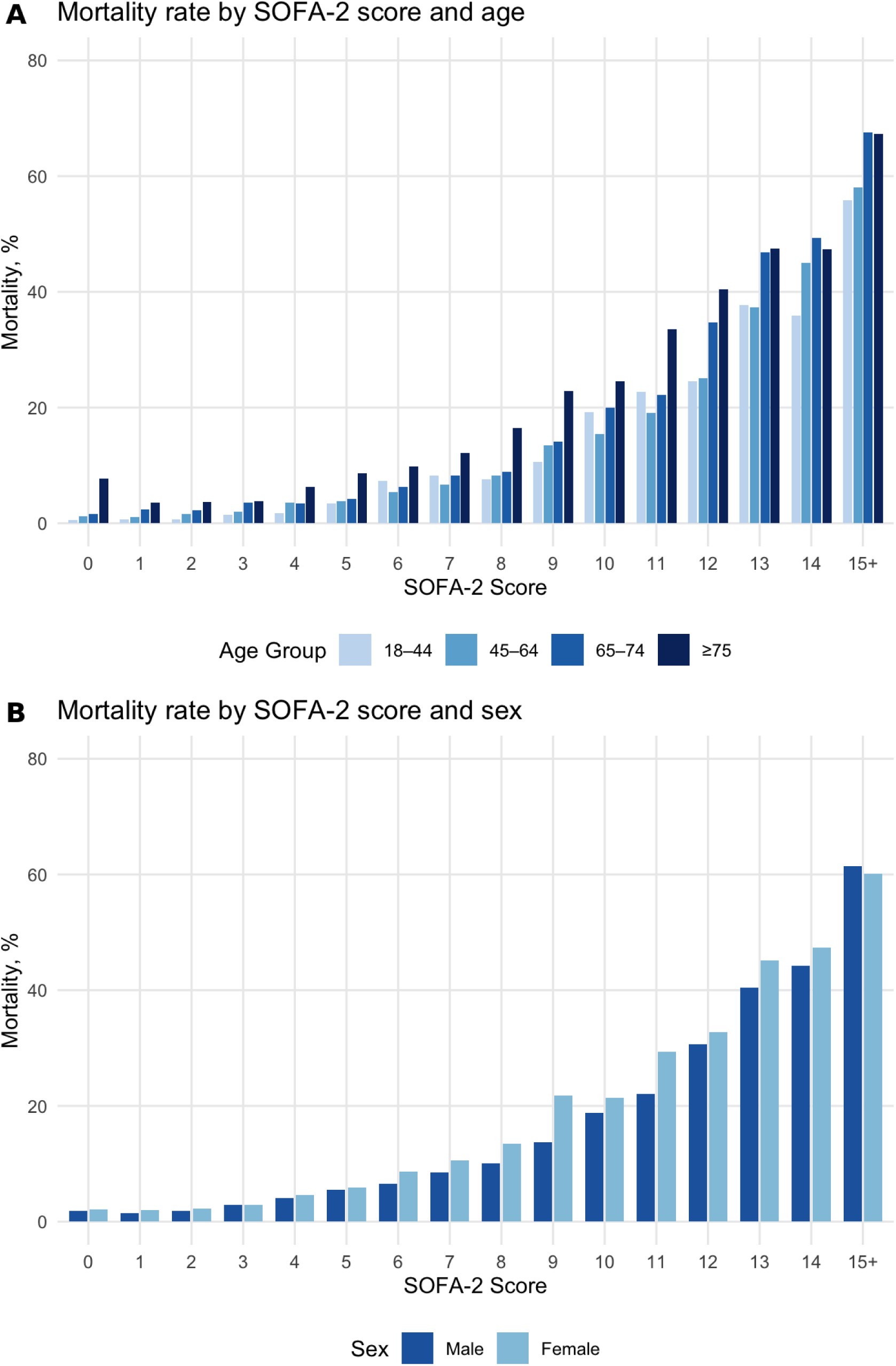
Observed ICU mortality by first-day SOFA-2 Score stratified by age and sex

**Table 2.**
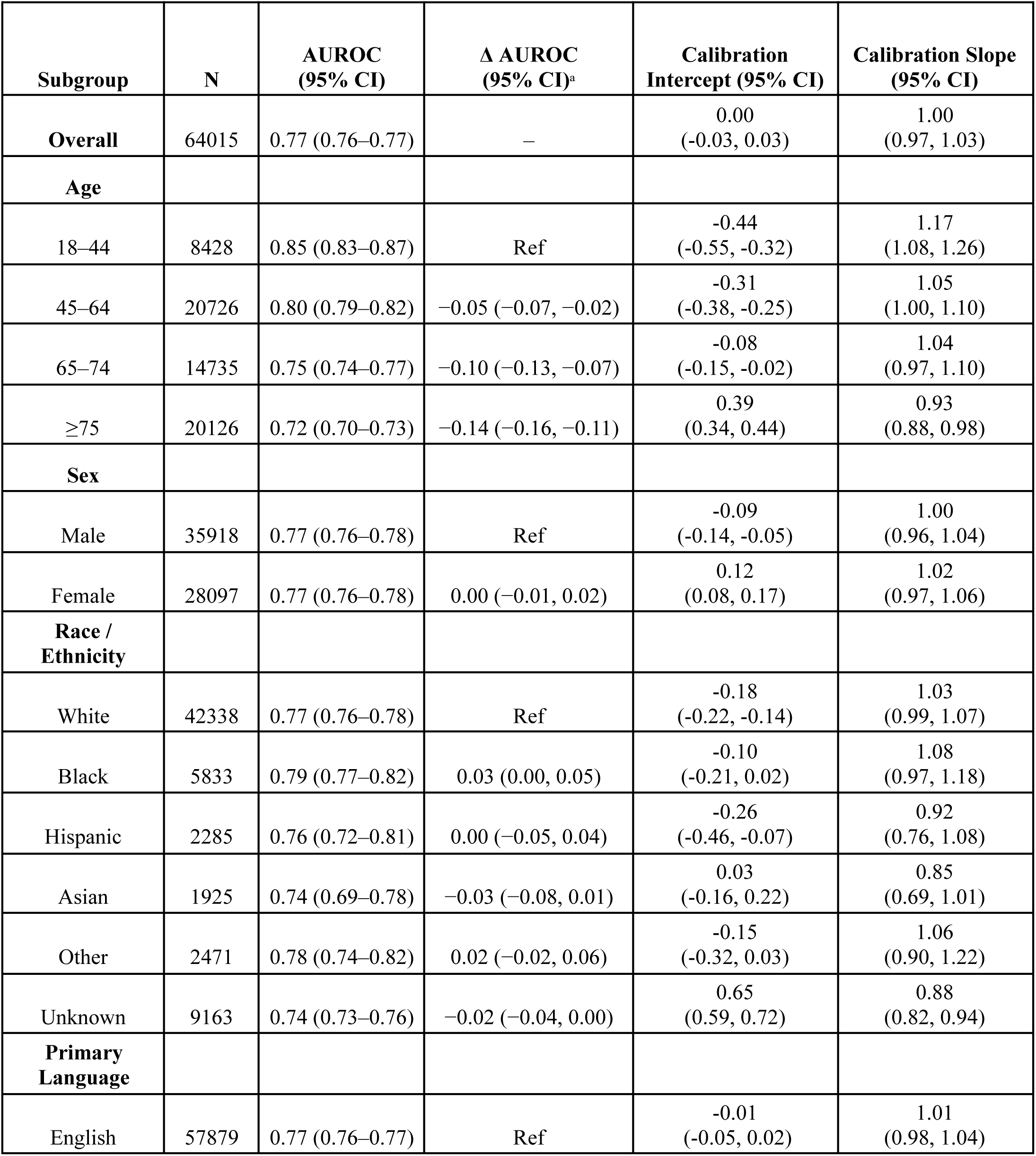
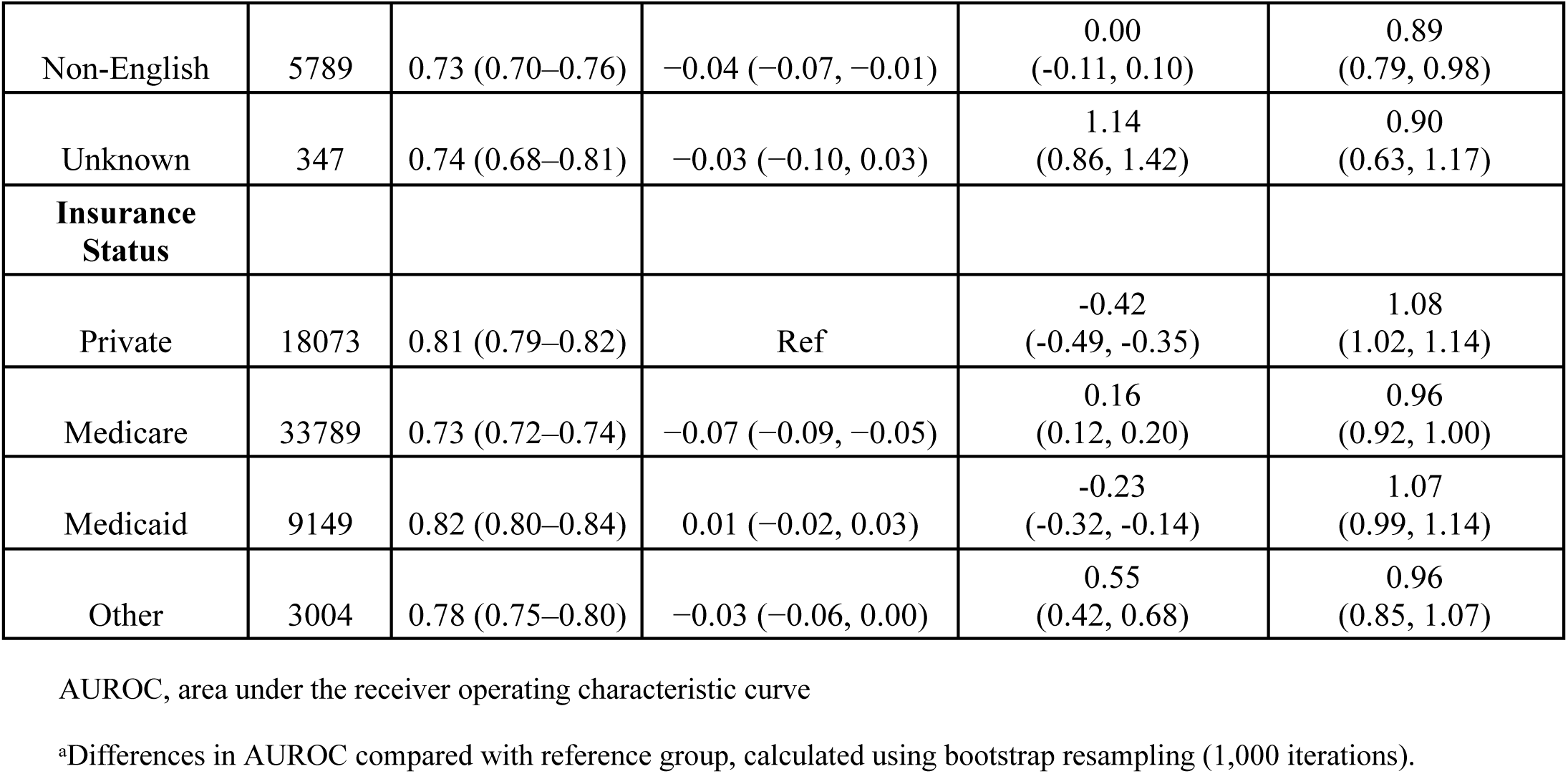
Discrimination and Calibration of SOFA-2 for ICU Mortality by Demographic Subgroup.

| Subgroup | N | AUROC<br>(95% CI) | $\Delta$ AUROC<br>(95% CI)<sup>a</sup> | Calibration<br>Intercept (95% CI) | Calibration Slope<br>(95% CI) |
| --- | --- | --- | --- | --- | --- |
| **Overall** | 64015 | 0.77 (0.76–0.77) | – | 0.00<br>(-0.03, 0.03) | 1.00<br>(0.97, 1.03) |
| **Age** |  |  |  |  |  |
| 18–44 | 8428 | 0.85 (0.83–0.87) | Ref | -0.44<br>(-0.55, -0.32) | 1.17<br>(1.08, 1.26) |
| 45–64 | 20726 | 0.80 (0.79–0.82) | -0.05 (-0.07, -0.02) | -0.31<br>(-0.38, -0.25) | 1.05<br>(1.00, 1.10) |
| 65–74 | 14735 | 0.75 (0.74–0.77) | -0.10 (-0.13, -0.07) | -0.08<br>(-0.15, -0.02) | 1.04<br>(0.97, 1.10) |
| ≥75 | 20126 | 0.72 (0.70–0.73) | -0.14 (-0.16, -0.11) | 0.39<br>(0.34, 0.44) | 0.93<br>(0.88, 0.98) |
| **Sex** |  |  |  |  |  |
| Male | 35918 | 0.77 (0.76–0.78) | Ref | -0.09<br>(-0.14, -0.05) | 1.00<br>(0.96, 1.04) |
| Female | 28097 | 0.77 (0.76–0.78) | 0.00 (-0.01, 0.02) | 0.12<br>(0.08, 0.17) | 1.02<br>(0.97, 1.06) |
| **Race /**<br>**Ethnicity** |  |  |  |  |  |
| White | 42338 | 0.77 (0.76–0.78) | Ref | -0.18<br>(-0.22, -0.14) | 1.03<br>(0.99, 1.07) |
| Black | 5833 | 0.79 (0.77–0.82) | 0.03 (0.00, 0.05) | -0.10<br>(-0.21, 0.02) | 1.08<br>(0.97, 1.18) |
| Hispanic | 2285 | 0.76 (0.72–0.81) | 0.00 (-0.05, 0.04) | -0.26<br>(-0.46, -0.07) | 0.92<br>(0.76, 1.08) |
| Asian | 1925 | 0.74 (0.69–0.78) | -0.03 (-0.08, 0.01) | 0.03<br>(-0.16, 0.22) | 0.85<br>(0.69, 1.01) |
| Other | 2471 | 0.78 (0.74–0.82) | 0.02 (-0.02, 0.06) | -0.15<br>(-0.32, 0.03) | 1.06<br>(0.90, 1.22) |
| Unknown | 9163 | 0.74 (0.73–0.76) | -0.02 (-0.04, 0.00) | 0.65<br>(0.59, 0.72) | 0.88<br>(0.82, 0.94) |
| **Primary**<br>**Language** |  |  |  |  |  |
| English | 57879 | 0.77 (0.76–0.77) | Ref | -0.01<br>(-0.05, 0.02) | 1.01<br>(0.98, 1.04) |
| Non-English | 5789 | 0.73 (0.70–0.76) | −0.04 (−0.07, −0.01) | 0.00<br>(−0.11, 0.10) | 0.89<br>(0.79, 0.98) |
| Unknown | 347 | 0.74 (0.68–0.81) | −0.03 (−0.10, 0.03) | 1.14<br>(0.86, 1.42) | 0.90<br>(0.63, 1.17) |
| **Insurance Status** |  |  |  |  |  |
| Private | 18073 | 0.81 (0.79–0.82) | Ref | −0.42<br>(−0.49, −0.35) | 1.08<br>(1.02, 1.14) |
| Medicare | 33789 | 0.73 (0.72–0.74) | −0.07 (−0.09, −0.05) | 0.16<br>(0.12, 0.20) | 0.96<br>(0.92, 1.00) |
| Medicaid | 9149 | 0.82 (0.80–0.84) | 0.01 (−0.02, 0.03) | −0.23<br>(−0.32, −0.14) | 1.07<br>(0.99, 1.14) |
| Other | 3004 | 0.78 (0.75–0.80) | −0.03 (−0.06, 0.00) | 0.55<br>(0.42, 0.68) | 0.96<br>(0.85, 1.07) |

AUROC, area under the receiver operating characteristic curve
<sup>a</sup>Differences in AUROC compared with reference group, calculated using bootstrap resampling (1,000 iterations).

### Performance across demographic categories

#### Age

Discrimination was highest among younger patients (age 18–44: AUROC 0.85; 95% CI: 0.83–0.87) and declined progressively with age, reaching its nadir in patients aged ≥75 (AUROC 0.72; 95% CI: 0.70–0.73). This decline of 0.14 (95% CI: 0.11–0.16) was statistically significant and represented the largest discrimination gap observed across any demographic category. Calibration exhibited a corresponding pattern: SOFA-2 overpredicted mortality among younger patients (intercept = −0.44; 95% CI: −0.55 to −0.32) and underpredicted mortality among the oldest group (intercept = 0.39; 95% CI: 0.34–0.44). This pattern is visually illustrated in Figure 2A, which shows that at any given SOFA-2 score, older patients experienced substantially higher observed mortality than younger patients. For example, at a SOFA-2 score of 10, mortality was 19.3% in patients aged 18–44 compared with 24.5% in those aged ≥75.

#### Sex

Performance was similar between sexes. Males and females had identical AUROCs of 0.77 (95% CI: 0.76–0.78). However, calibration differed between sexes, with SOFA-2 slightly overpredicting mortality in males (intercept = −0.09) and underpredicting in females (intercept = 0.12). Figure 2B demonstrates that for equivalent SOFA-2 scores, observed mortality was generally higher among females, with convergence at scores ≥15 where both groups approached 61% mortality.

### Race/Ethnicity

Among patients with documented race and ethnicity, discrimination was consistent across groups, with AUROCs ranging from 0.74 (Asian) to 0.79 (Black); no differences were statistically significant compared to the White reference group (AUROC 0.77). Calibration was also similar, with intercepts ranging from −0.26 (Hispanic) to 0.03 (Asian), indicating modest variation across documented racial and ethnic groups.

However, patients with unknown race/ethnicity (14.3% of the cohort) demonstrated notably different characteristics. This group had the highest observed ICU mortality rate (14.1%, nearly double the overall rate of 7.2%), longer median ICU lengths of stay (2.2 days [IQR, 1.2–4.9] vs approximately 1.9 days for documented groups), and poorer calibration (intercept = 0.65; 95% CI: 0.59–0.72), indicating systematic underprediction of mortality in this subgroup.

### Language

Discrimination was significantly lower for non-English speakers (AUROC 0.73; 95% CI: 0.70–0.76) compared to English speakers (AUROC 0.77; ΔAUROC −0.04; 95% CI: −0.07 to −0.01). Calibration was similar between these two groups. Patients with Unknown language status, though representing only 0.5% of the cohort (n=347), had the highest ICU mortality rate observed in the study (23.1%), longer median ICU length of stay (2.1 days [IQR, 1.1–5.1]), and substantially poorer calibration (intercept = 1.14; 95% CI: 0.86–1.42).

### Insurance Status

Discrimination varied substantially by insurance status, with the highest performance observed among patients with Medicaid (AUROC 0.82; 95% CI: 0.80–0.84) and private insurance (AUROC 0.81; 95% CI: 0.79–0.82), and significantly lower performance among Medicare recipients (AUROC 0.73; 95% CI: 0.72–0.74; ΔAUROC −0.07; 95% CI: −0.09 to −0.05). Calibration reflected these patterns: SOFA-2 overpredicted mortality for private (intercept = −0.42) and Medicaid (intercept = −0.23) patients while underpredicting for Medicare (intercept = 0.16) and other insurance (intercept = 0.55) patients.

## Discussion

In this external validation study of over 64,000 ICU admissions from MIMIC-IV, SOFA-2 demonstrated good overall discrimination and calibration for predicting ICU mortality, consistent with the original development cohort. However, we identified clinically meaningful variation in performance across demographic subgroups.

The most notable finding was the progressive decline in discrimination with advancing age, with a ΔAUROC of −0.14 between the youngest and oldest patient groups. This was accompanied by systematic underprediction of mortality in older adults. Relatedly, discrimination was significantly lower among Medicare recipients compared to those with private insurance and Medicaid, a pattern likely driven by age given Medicare eligibility.

These findings align with previous literature demonstrating limited accuracy of the SOFA score in older adults.^19^ Given that patients aged ≥75 constitute nearly one-third of ICU admissions in this dataset and represent a growing proportion of critically ill patients nationally, this finding has direct implications for triage, prognostication, and goals-of-care discussions.^20,21^ The poorer performance of SOFA-2 among older adults could reflect limitations in what acute physiological parameters can capture when comorbidity burden and diminished physiological reserve all contribute to outcomes, information SOFA-2 does not capture.^22,23^ Prior work combining a metric of frailty with the SOFA score did not meaningfully improve performance among older adults, however,^19^ suggesting that other unmeasured factors may be playing a role, such as increased risk of ICU delirium among older adults with dementia,^19^ or decisions about withdrawal of life-sustaining care that impact ICU survival.

The significantly lower discrimination among non-English speakers may reflect systematic differences in clinical documentation across language barriers,^13^ unmeasured social determinants, or differences in provision of care. While smaller than the age-related gap, this finding warrants further investigation given prior associations between language-concordant care and improved outcomes.^24^

Our analysis also revealed a concerning pattern among patients with missing demographic information.^25,26^ Patients with Unknown race/ethnicity (14.3% of the cohort) experienced nearly double the overall mortality rate (14.1% vs. 7.2%), while those with unknown language status had the highest mortality observed (23.1%). Both groups had longer ICU stays and poor calibration, suggesting that missing data may mark unmeasured factors associated with poor outcomes, such as acute illness severity precluding complete documentation or systematic underrepresentation of vulnerable populations. This has implications both for clinical interpretation and future fairness research, where excluding patients with missing data may serve to remove the highest-risk individuals.

These findings extend prior work on the original SOFA score. Multiple studies have shown SOFA overestimates mortality among Black patients, potentially disadvantaging them in crisis standards of care allocation, attributed partly to racial variation in baseline creatinine.^4–6,12,27^ In our study, discrimination did not differ significantly across racial and ethnic groups with documented status, though the high proportion of patients with Unknown race/ethnicity limits interpretation. Sex-based differences have also been reported previously, with women demonstrating lower SOFA scores that do not correspond to improved outcomes.^8^ We observed a similar pattern: while discrimination was equivalent between sexes, SOFA-2 modestly overpredicted mortality in males (intercept −0.09) and underpredicted in females (intercept 0.12), consistent with prior findings that equivalent scores carry different prognostic weight by sex.

These results suggest interpreting SOFA-2 with greater caution in older adult patients, recognizing that equivalent scores may carry different prognostic implications by sex, attending to potential documentation differences for non-English speakers,^28^ and treating incomplete demographic information as a possible signal of higher baseline risk. While SOFA-2 updated organ dysfunction thresholds using data from over 3 million admissions across 9 countries, it did not include systematic subgroup evaluation, potentially carrying forward unexamined sources of bias. These findings underscore the importance of routine equity evaluation as a complement to traditional validation of clinical prediction tools.

## Limitations

This study has several limitations. Our analysis used data from a single academic medical center in the northeastern United States, which may limit generalizability to other settings. We relied on administratively recorded race/ethnicity categories, which may not reflect patient self-identification and are subject to misclassification, as highlighted by the proportion of patients with Unknown race/ethnicity (14.3%). Several SOFA-2 components could not be fully abstracted from the EHR, including machine availability constraints, nonrenal indications for renal replacement therapy, and treatment ceilings.^29^ Pre-sedation neurologic assessment also was not incorporated because sedation timing relative to GCS measurement could not be reliably determined. Further, we did not account for advance care planning, surrogate decision-making, or withdrawal of life-sustaining treatment, which could have differentially impacted outcomes across subgroups. Selection bias may reflect inclusion criteria, as older adults are more frequently rejected for ICU transfer,^30^ so our results only reflect performance among admitted patients. Finally, we assessed first-day SOFA-2 scores only and did not evaluate whether fairness properties differ with serial score trajectories.

## Conclusions

In this external validation study, SOFA-2 demonstrated good overall performance for ICU mortality prediction, but performance varied meaningfully across demographic subgroups. Discrimination declined substantially with advancing age and was lower among patients whose primary language was not English, while patients with missing demographic information showed particularly poor calibration and higher observed mortality. These findings suggest that routine equity evaluation should complement traditional validation of clinical prediction tools before widespread implementation.

## Data Availability

The deidentified individual patient data underlying this study are available through the MIMIC-IV database (version 3.1), hosted on PhysioNet. MIMIC-IV is publicly available to credentialed users who complete required human-subjects training through the CITI program and sign a data use agreement. The complete analytic code (SQL queries and R scripts) is publicly available at https://github.com/SichengH/SOFA2_bias.

https://physionet.org/content/mimiciv/3.1/

## Data Sharing Statement

**Data:** The deidentified individual patient data underlying this study are available through the MIMIC-IV database (version 3.1), hosted on PhysioNet (https://physionet.org/content/mimiciv/3.1/).

**Available Data:** Individual-level data, a data dictionary, and the complete analytic code (SQL queries and R scripts) used to generate study results.

**Data Access:** MIMIC-IV is publicly available to credentialed users who complete required human-subjects training through the Collaborative Institutional Training Initiative (CITI) program and sign a data use agreement via PhysioNet. There are no additional restrictions on the types of analyses that may be performed. Analytic code is publicly available at https://github.com/SichengH/SOFA2_bias.

## Ethics Statement

This study used deidentified, publicly available data from the MIMIC-IV database hosted on PhysioNet. All authors completed required human-subjects training through the CITI program and signed a data use agreement. No institutional review board approval was required.

## References

1. Vincent JL, Moreno R, Takala J, et al. The SOFA (Sepsis-related Organ Failure Assessment) score to describe organ dysfunction/failure. On behalf of the Working Group on Sepsis-Related Problems of the European Society of Intensive Care Medicine. Intensive Care Med. 1996;22(7):707–710. doi:10.1007/BF01709751

2. Raith EP, Udy AA, Bailey M, et al. Prognostic Accuracy of the SOFA Score, SIRS Criteria, and qSOFA Score for In-Hospital Mortality Among Adults With Suspected Infection Admitted to the Intensive Care Unit. JAMA. 2017;317(3):290–300. doi:10.1001/jama.2016.20328

3. Singer M, Deutschman CS, Seymour CW, et al. The Third International Consensus Definitions for Sepsis and Septic Shock (Sepsis-3). JAMA. 2016;315(8):801–810. doi:10.1001/jama.2016.0287

4. Miller WD, Han X, Peek ME, Charan Ashana D, Parker WF. Accuracy of the Sequential Organ Failure Assessment Score for In-Hospital Mortality by Race and Relevance to Crisis Standards of Care. JAMA Netw Open. 2021;4(6):e2113891. doi:10.1001/jamanetworkopen.2021.13891

5. Tolchin B, Oladele C, Galusha D, et al. Racial disparities in the SOFA score among patients hospitalized with COVID-19. PLoS ONE. 2021;16(9):e0257608. doi:10.1371/journal.pone.0257608

6. Ashana DC, Anesi GL, Liu VX, et al. Equitably Allocating Resources during Crises: Racial Differences in Mortality Prediction Models. Am J Respir Crit Care Med. 2021;204(2):178–186. doi:10.1164/rccm.202012-4383OC

7. White DB, Lo B. Mitigating Inequities and Saving Lives with ICU Triage during the COVID-19 Pandemic. Am J Respir Crit Care Med. 2021;203(3):287–295. doi:10.1164/rccm.202010-3809CP

8. Zimmermann T, Kaufmann P, Amacher SA, et al. Sex differences in the SOFA score of ICU patients with sepsis or septic shock: a nationwide analysis. Crit Care. 2024;28(1):209. doi:10.1186/s13054-024-04996-y

9. Ranzani OT, Singer M, Salluh JIF, et al. Development and Validation of the Sequential Organ Failure Assessment (SOFA)-2 Score. JAMA. Published online October 29, 2025. doi:10.1001/jama.2025.20516

10. Riviello ED, Dechen T, O’Donoghue AL, et al. Assessment of a Crisis Standards of Care Scoring System for Resource Prioritization and Estimated Excess Mortality by Race, Ethnicity, and Socially Vulnerable Area During a Regional Surge in COVID-19. JAMA Netw Open. 2022;5(3):e221744. doi:10.1001/jamanetworkopen.2022.1744

11. Johnson AEW, Bulgarelli L, Shen L, et al. MIMIC-IV, a freely accessible electronic health record dataset. Sci Data. 2023;10(1):1. doi:10.1038/s41597-022-01899-x

12. Hermsen M, Lyons PG, Persad G, et al. Age and Saving Lives in Crisis Standards of Care: A Multicenter Cohort Study of Triage Score Prognostic Accuracy. Crit Care Explor. 2025;7(5):e1256. doi:10.1097/CCE.0000000000001256

13. Divi C, Koss RG, Schmaltz SP, Loeb JM. Language proficiency and adverse events in US hospitals: a pilot study. Int J Qual Health Care J Int Soc Qual Health Care. 2007;19(2):60–67. doi:10.1093/intqhc/mzl069

14. Fowler RA, Noyahr LA, Thornton JD, et al. An official American Thoracic Society systematic review: the association between health insurance status and access, care delivery, and outcomes for patients who are critically ill. Am J Respir Crit Care Med. 2010;181(9):1003–1011. doi:10.1164/rccm.200902-0281ST

15. Mandrekar JN. Receiver operating characteristic curve in diagnostic test assessment. J Thorac Oncol Off Publ Int Assoc Study Lung Cancer. 2010;5(9):1315–1316. doi:10.1097/JTO.0b013e3181ec173d

16. Steyerberg EW, Vickers AJ, Cook NR, et al. Assessing the performance of prediction models: a framework for traditional and novel measures. Epidemiology. 2010;21(1):128–138. doi:10.1097/EDE.0b013e3181c30fb2

17. Collins GS, Moons KGM, Dhiman P, et al. TRIPOD+AI statement: updated guidance for reporting clinical prediction models that use regression or machine learning methods. BMJ. 2024;385:e078378. doi:10.1136/bmj-2023-078378

18. Goldberger AL, Amaral LA, Glass L, et al. PhysioBank, PhysioToolkit, and PhysioNet: components of a new research resource for complex physiologic signals. Circulation. 2000;101(23):E215–220. doi:10.1161/01.cir.101.23.e215

19. Langlais E, Nesseler N, Le Pabic E, Frasca D, Launey Y, Seguin P. Does the clinical frailty score improve the accuracy of the SOFA score in predicting hospital mortality in elderly critically ill patients? A prospective observational study. J Crit Care. 2018;46:67–72. doi:10.1016/j.jcrc.2018.04.012

20. Hansen AV, Mortensen LH, Ekstrøm CT, Trompet S, Westendorp R. Predicting mortality and visualizing health care spending by predicted mortality in Danes over age 65. Sci Rep. 2023;13(1):1203. doi:10.1038/s41598-023-28102-4

21. Silva Júnior JM, Chaves RC de F, Corrêa TD, et al. Epidemiology and outcome of high-surgical-risk patients admitted to an intensive care unit in Brazil. Rev Bras Ter Intensiva. 2020;32(1):17–27. doi:10.5935/0103-507x.20200005

22. Zampieri FG, Colombari F. The impact of performance status and comorbidities on the short-term prognosis of very elderly patients admitted to the ICU. BMC Anesthesiol. 2014;14:59. doi:10.1186/1471-2253-14-59

23. Minne L, Ludikhuize J, de Jonge E, de Rooij S, Abu-Hanna A. Prognostic models for predicting mortality in elderly ICU patients: a systematic review. Intensive Care Med. 2011;37(8):1258–1268. doi:10.1007/s00134-011-2265-6

24. Diamond L, Izquierdo K, Canfield D, Matsoukas K, Gany F. A Systematic Review of the Impact of Patient–Physician Non-English Language Concordance on Quality of Care and Outcomes. J Gen Intern Med. 2019;34(8):1591–1606. doi:10.1007/s11606-019-04847-5

25. Sauer CM, Chen LC, Hyland SL, Girbes A, Elbers P, Celi LA. Leveraging electronic health records for data science: common pitfalls and how to avoid them. Lancet Digit Health. 2022;4(12):e893–e898. doi:10.1016/S2589-7500(22)00154-6

26. Casey JA, Schwartz BS, Stewart WF, Adler NE. Using Electronic Health Records for Population Health Research: A Review of Methods and Applications. Annu Rev Public Health. 2016;37:61–81. doi:10.1146/annurev-publhealth-032315-021353

27. Jamali H, Castillo LT, Morgan CC, et al. Racial Disparity in Oxygen Saturation Measurements by Pulse Oximetry: Evidence and Implications. Ann Am Thorac Soc. 2022;19(12):1951–1964. doi:10.1513/AnnalsATS.202203-270CME

28. Limaye NP, Matias WR, Rozansky H, et al. Limited English Proficiency and Sepsis Mortality by Race and Ethnicity. JAMA Netw Open. 2024;7(1):e2350373. doi:10.1001/jamanetworkopen.2023.50373

29. Uchino S, Katayama S. Revisiting the simplicity of SOFA-2: a pragmatic assessment of implementability. Crit Care. 2026;30:82. doi:10.1186/s13054-026-05892-3

30. Sprung CL, Artigas A, Kesecioglu J, et al. The Eldicus prospective, observational study of triage decision making in European intensive care units. Part II: Intensive care benefit for the elderly. Crit Care Med. 2012;40(1):132. doi:10.1097/CCM.0b013e318232d6b0

31. Austin PC, Stuart EA. Moving towards best practice when using inverse probability of treatment weighting (IPTW) using the propensity score to estimate causal treatment effects in observational studies. Stat Med. 2015;34(28):3661–3679. doi:10.1002/sim.6607

