## Supplemental Material for "Evaluation of SOFA-2 Score Performance Across Demographic Subgroups: An External Validation Study Using MIMIC-IV"

### TRIPOD-AI Checklist

| Section/Topic | Item | Checklist item | Reported on page/section |
|---|---|---|---|
| **TITLE** | | | |
| Title | 1 | Identify the study as developing or evaluating the performance of a multivariable prediction model, the target population, and the outcome to be predicted | Title page. Title identifies evaluation of SOFA-2, target population, and outcome. |
| **ABSTRACT** | | | |
| Abstract | 2 | See TRIPOD+AI for Abstracts checklist | Abstract. Structured abstract addresses TRIPOD+AI for Abstracts items, except that the study was not registered (item 13). |
| **INTRODUCTION** | | | |
| Background | 3a | Explain the healthcare context (including whether diagnostic or prognostic) and rationale for developing or evaluating the prediction model, including references to existing models | Introduction, paragraphs 1–2. Prognostic context; references to original SOFA and prior fairness studies. |
| | 3b | Describe the target population and the intended purpose of the prediction model in the context of the care pathway, including its intended users (e.g., healthcare professionals, patients, public) | Introduction, paragraphs 2–3. ICU patients; SOFA-2 used for triage, prognostication, research and clinical decision-making. |
| | 3c | Describe any known health inequalities between sociodemographic groups | Introduction, paragraph 2. Racial/ethnic disparities in SOFA performance, sex differences, crisis care implications. |
| Objectives | 4 | Specify the study objectives, including whether the study describes the development or validation of a prediction model (or both) | Introduction, paragraph 4. External validation of SOFA-2. |
| **METHODS** | | | |
| Data | 5a | Describe the sources of data separately for the development and evaluation datasets (e.g., randomised trial, cohort, routine care or registry data), the rationale for using these data, and representativeness of the data | Methods, Study design and data source. MIMIC-IV v3.1; rationale: external to SOFA-2 development cohort. |
| | 5b | Specify the dates of the collected participant data, including start and end of participant accrual; and, if applicable, end of follow-up | Methods, Study design and data source. 2008–2022. |
| Participants | 6a | Specify key elements of the study setting (e.g., primary care, secondary care, general population) including the number and location of centres | Methods, Study design and data source. Single tertiary care center: Beth Israel Deaconess Medical Center. |
| | 6b | Describe the eligibility criteria for study participants | Methods, Study population. Adults ≥18 years, first ICU admission, excluded if LOS <6 hours or implausible physiologic values (eTable 1). |
| | 6c | Give details of any treatments received, and how they were handled during model development or evaluation, if relevant | Methods, SOFA-2 Score calculation. |

| | | | |
|---|---|---|---|
| Data preparation | 7 | Describe any data pre-processing and quality checking, including whether this was similar across relevant sociodemographic groups | Methods, Study population and SOFA-2 Score calculation. Implausible value exclusion (eTable 1); Fig. 1 monitors demographic shifts at each exclusion step. |
| Outcome | 8a | Clearly define the outcome that is being predicted and the time horizon, including how and when assessed, the rationale for choosing this outcome, and whether the method of outcome assessment is consistent across sociodemographic groups | Methods, Outcome. ICU mortality. Chosen for comparability with prior work. Assessment method uniform across subgroups. |
| | 8b | If outcome assessment requires subjective interpretation, describe the qualifications and demographic characteristics of the outcome assessors | N/A. ICU mortality is an objective outcome recorded in the EHR. |
| | 8c | Report any actions to blind assessment of the outcome to be predicted | N/A. Retrospective study using EHR data; outcome is objective (death). |
| Predictors | 9a | Describe the choice of initial predictors (e.g., literature, previous models, all available predictors) and any pre-selection of predictors before model building | N/A (evaluation only). SOFA-2 predictors predefined by Ranzani et al. |
| | 9b | Clearly define all predictors, including how and when they were measured (and any actions to blind assessment of predictors for the outcome and other predictors) | Methods, SOFA-2 Score calculation. Six organ systems defined; worst values in first 24 hours; specific thresholds and modalities described. |
| | 9c | If predictor measurement requires subjective interpretation, describe the qualifications and demographic characteristics of the predictor assessors | Methods, SOFA-2 Score calculation. Clinical data recorded by clinicians. |
| Sample size | 10 | Explain how the study size was arrived at (separately for development and evaluation), and justify that the study size was sufficient to answer the research question. Include details of sample size calculation | Methods, Study population / Results. All eligible MIMIC-IV patients included (n=64,015). Full-cohort analysis. |
| Missing data | 11 | Describe how missing data were handled. Provide reasons for omitting any data | Methods, SOFA-2 Score calculation; eTable 2. Missing components assigned score of 0 (normal function), consistent with original SOFA-2 validation. Missingness rates reported by component. |
| Analytical methods | 12a | Describe how the data were used (e.g., for development and evaluation of model performance) in the analysis, including whether the data were partitioned, considering any sample size requirements | N/A (evaluation only). |
| | 12b | Depending on the type of model, describe how predictors were handled in the analyses (functional form, rescaling, transformation, or any standardisation) | N/A (evaluation only). SOFA-2 total score used as sole predictor per original specification. |
| | 12c | Specify the type of model, rationale, all model-building steps, including any hyperparameter tuning, and method for internal validation | N/A (evaluation only). |
| | 12d | Describe if and how any heterogeneity in estimates of model parameter values and model performance was handled and quantified across clusters (e.g., hospitals, countries) | Methods, Statistical analysis. Single-center study. Subgroup heterogeneity assessed via bootstrap AUROC differences with 95% CIs. |
| | 12e | Specify all measures and plots used (and their rationale) to evaluate model performance (e.g., discrimination, calibration, clinical utility) and, if relevant, to compare multiple models | Methods, Statistical analysis. AUROC (discrimination), calibration intercepts and slopes; calibration plots (eFigures 2–6); observed mortality by score (Figure 2, eFigures 7–9). AUROC benchmarks cited. |

| | | | |
|---|---|---|---|
| | 12f | Describe any model updating (e.g., recalibration) arising from the model evaluation, either overall or for particular sociodemographic groups or settings | No model updating was performed. This study assessed existing SOFA-2 performance without recalibration. |
| | 12g | For model evaluation, describe how the model predictions were calculated (e.g., formula, code, object, application programming interface) | Methods, SOFA-2 Score calculation. Predictions from logistic regression with SOFA-2 as sole predictor on full cohort. All code publicly available at GitHub repository. |
| Class imbalance | 13 | If class imbalance methods were used, state why and how this was done, and any subsequent methods to recalibrate the model or the model predictions | No class imbalance methods were used. |
| Fairness | 14 | Describe any approaches that were used to address model fairness and their rationale | Introduction, paragraph 4; Methods, Demographic subgroups and Statistical analysis. Fairness operationalized as equivalence in discrimination and calibration across subgroups. |
| Model output | 15 | Specify the output of the prediction model (e.g., probabilities, classification). Provide details and rationale for any classification and how the thresholds were identified | N/A (evaluation only). SOFA-2 outputs an integer score (0–24); logistic regression generated predicted probabilities for calibration assessment. |
| Training versus evaluation | 16 | Identify any differences between the development and evaluation data in healthcare setting, eligibility criteria, outcome, and predictors | Methods, Study design; Discussion, Limitations. MIMIC-IV (single US academic center, 2008–2022) is external to development data. Same outcome and predictor. Differences in setting acknowledged. |
| Ethical approval | 17 | Name the institutional research board or ethics committee that approved the study and describe the participant-informed consent or the ethics committee waiver of informed consent | Methods, Ethics and approval. Deidentified, publicly available data from PhysioNet; CITI training and DUA required. No IRB approval needed. |
| **OPEN SCIENCE** | | | |
| Funding | 18a | Give the source of funding and the role of the funders for the present study | COI/funding section. |
| Conflicts of interest | 18b | Declare any conflicts of interest and financial disclosures for all authors | COI/funding section. |
| Protocol | 18c | Indicate where the study protocol can be accessed or state that a protocol was not prepared | No formal study protocol was prepared. |
| Registration | 18d | Provide registration information for the study, including register name and registration number, or state that the study was not registered | This study was not registered. It is a retrospective external validation using publicly available data. |
| Data sharing | 18e | Provide details of the availability of the study data | Data Sharing Statement. MIMIC-IV v3.1 on PhysioNet. Available to credentialed users. |
| Code sharing | 18f | Provide details of the availability of the analytical code | Data Sharing Statement; Methods, Statistical analysis. Complete SQL and R code at https://github.com/SichengH/SOFA2_bias. |
| **PATIENT & PUBLIC INVOLVEMENT** | | | |
| Patient & Public Involvement | 19 | Provide details of any patient and public involvement during the design, conduct, reporting, interpretation, or dissemination of the study or state no involvement | There was no patient or public involvement in the design, conduct, or reporting of this study. |

| | | | |
|---|---|---|---|
| **RESULTS** | | | |
| Participants | 20a | Describe the flow of participants through the study, including the number of participants with and without the outcome and, if applicable, a summary of the follow-up time. A diagram may be helpful | Results, paragraph 1; Figure 1. |
| | 20b | Report the characteristics overall and, where applicable, for each data source or setting, including the key dates, key predictors (including demographics), treatments received, sample size, number of outcome events, follow-up time, and amount of missing data. Report any differences across key demographic groups | Results, Cohort characteristics; Table 1; eTable 2; eTable 3. Characteristics by demographic subgroup. Missingness by SOFA-2 component. Organ subscores by subgroup. |
| | 20c | For model evaluation, show a comparison with the development data of the distribution of important predictors (demographics, predictors, and outcome) | Results, Cohort characteristics and Overall SOFA-2 performance. Direct predictor-level comparison limited by aggregated reporting in original study. |
| Model development | 21 | Specify the number of participants and outcome events in each analysis (e.g., for model development, hyperparameter tuning, model evaluation) | Results. n=64,015 with 4,596 outcome events. Subgroup-specific n and outcome counts in Table 1 and Table 2. |
| Model specification | 22 | Provide details of the full prediction model (e.g., formula, code, object, application programming interface) to allow predictions in new individuals and to enable third-party evaluation and implementation | N/A (evaluation only). SOFA-2 specification per Ranzani et al. Implementation code available on GitHub. |
| Model performance | 23a | Report model performance estimates with confidence intervals, including for any key subgroups (e.g., sociodemographic). Consider plots to aid presentation | Results, Overall SOFA-2 performance and Performance across demographic categories. AUROC, calibration and 95% CIs for subgroups. |
| | 23b | If examined, report results of any heterogeneity in model performance across clusters | Subgroup heterogeneity reported via bootstrap AUROC differences |
| Model updating | 24 | Report the results from any model updating, including the updated model and subsequent performance | No model updating was performed. |
| **DISCUSSION** | | | |
| Interpretation | 25 | Give an overall interpretation of the main results, including issues of fairness in the context of the objectives and previous studies | Discussion. Age-related decline in discrimination interpreted in context of prior SOFA fairness literature; language and missing data findings discussed with clinical implications. |
| Limitations | 26 | Discuss any limitations of the study (such as a non-representative sample, sample size, overfitting, missing data) and their effects on any biases, statistical uncertainty, and generalizability | Limitations. Single center, administrative race/ethnicity, incomplete EHR data, no ACP data, selection bias in ICU admission, first-day scores only. |
| Usability of the model in the context of current care | 27a | Describe how poor quality or unavailable input data (e.g., predictor values) should be assessed and handled when implementing the prediction model | N/A (evaluation only). However, Methods and Limitations note missing component handling (score of 0) and EHR abstraction challenges. |
| | 27b | Specify whether users will be required to interact in the handling of the input data or use of the model, and what level of expertise is required of users | N/A (evaluation only). |
| | 27c | Discuss any next steps for future research, with a specific view to applicability and generalizability of the model | Discussion, final paragraph and Conclusions. Calls for routine equity evaluation of clinical prediction tools. |

**eTable 1.** Acceptable ranges of physiologic variables employed in this study

| Variable | Acceptable Range of Values |
|---|---|
| Glasgow Coma Scale | 3–15 |
| Glasgow Coma Scale: Motor component | 1–6 |
| Platelets | ≥1, ≤999 (x10^3/µL) |
| Total Serum Bilirubin | ≥1.70, ≤800 µmol/L<br>(≥0.1, ≤46.78 mg/dl) |
| Mean Arterial Pressure (MAP) | ≥15, ≤299 mmHg |
| Serum Creatinine | ≥8.8, ≤999 µmol/L<br>(≥0.10, ≤11.23 mg/dl) |

**eTable 2:** Missingness of SOFA-2 Components and Variables During the First 24 Hours of ICU Admission

| **Variable** | **MIMIC-IV (n=64015)** |
|---|---|
| | n (%) Missing |
| **SOFA-2 Components** | |
| Neurological | 141 (0.2%) |
| Hepatic | 37114 (58.0%) |
| Cardiovascular | 0 (0.0%) |
| Renal | 730 (1.1%) |
| Respiratory | 41450 (64.8%) |
| Coagulation | 2605 (4.1%) |
| Complete SOFA-2 (all 6 components) | 8949 (14.0%) |
| Complete SOFA-2 (excl. respiratory) | 26699 (41.7%) |

*Missingness at the component level indicates that none of the underlying physiologic variables for a given organ system were recorded during the first 24 hours of ICU admission. This may reflect true data unavailability or, more commonly, the absence of clinically indicated testing (e.g., bilirubin is typically measured only when hepatic dysfunction is suspected, accounting for the 58.0% missingness rate). Consistent with the original SOFA-2 validation, missing components were assigned a score of 0 (normal function).*

**eFigure 1:** Distribution of first-day SOFA-2 Scores in the study population (n=64015)

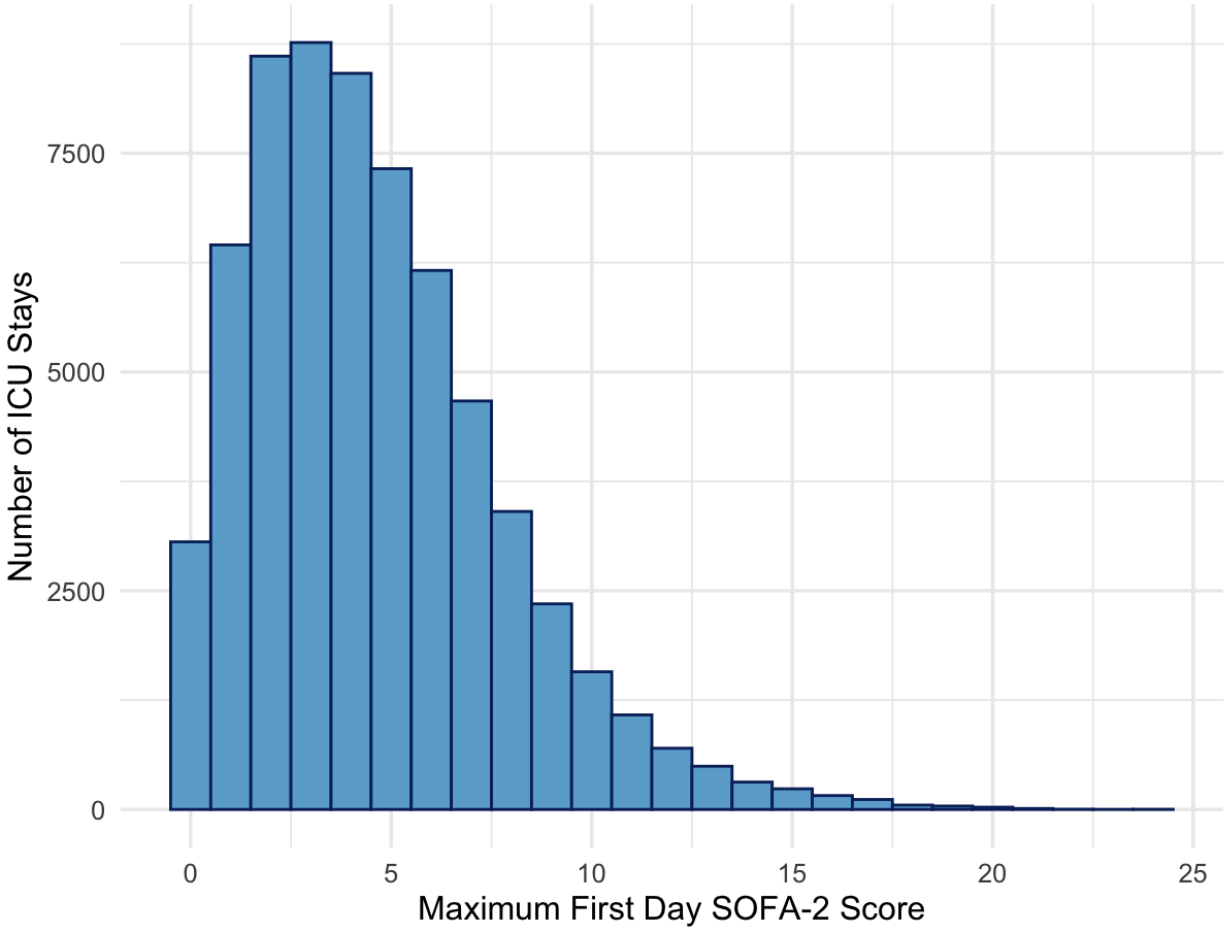

**eTable 3:** Median SOFA-2 Organ-Specific Subscores by Demographic Subgroup

| Subgroup | N | Neuro | Cardio | Respiratory | Hepatic | Renal | Coag | Median SOFA2 |
|---|---|---|---|---|---|---|---|---|
| **Overall** | 64015 | 0 (0–1) | 2 (1–2) | 0 (0–1) | 0 (0–0) | 1 (0–1) | 0 (0–1) | 4 (2–6) |
| **Sex** | | | | | | | | |
| Male | 35918 | 0 (0–1) | 2 (1–2) | 0 (0–1) | 0 (0–0) | 1 (0–1) | 0 (0–1) | 4 (2–7) |
| Female | 28097 | 0 (0–1) | 2 (1–2) | 0 (0–0) | 0 (0–0) | 1 (0–1) | 0 (0–1) | 4 (2–6) |
| **Age** | | | | | | | | |
| 18–44 | 8428 | 0 (0–1) | 1 (0–2) | 0 (0–0) | 0 (0–0) | 0 (0–1) | 0 (0–1) | 3 (1–5) |
| 45–64 | 20726 | 0 (0–1) | 1 (0–2) | 0 (0–1) | 0 (0–0) | 1 (0–1) | 0 (0–1) | 4 (2–6) |
| 65–74 | 14735 | 0 (0–1) | 2 (1–2) | 0 (0–1) | 0 (0–0) | 1 (0–1) | 0 (0–1) | 4 (3–7) |
| ≥75 | 20126 | 1 (0–1) | 2 (1–2) | 0 (0–1) | 0 (0–0) | 1 (0–2) | 0 (0–1) | 5 (3–7) |
| **Race/Ethnicity** | | | | | | | | |
| White | 42338 | 0 (0–1) | 2 (1–2) | 0 (0–1) | 0 (0–0) | 1 (0–1) | 0 (0–1) | 4 (2–6) |
| Black | 5833 | 0 (0–1) | 1 (0–2) | 0 (0–0) | 0 (0–0) | 1 (0–2) | 0 (0–1) | 4 (2–6) |
| Hispanic | 2285 | 0 (0–1) | 1 (0–2) | 0 (0–0) | 0 (0–0) | 1 (0–1) | 0 (0–1) | 4 (2–6) |
| Asian | 1925 | 0 (0–1) | 1 (1–2) | 0 (0–0) | 0 (0–0) | 0 (0–1) | 0 (0–1) | 4 (2–6) |
| Other | 2471 | 0 (0–1) | 1 (1–2) | 0 (0–1) | 0 (0–0) | 1 (0–1) | 0 (0–1) | 4 (2–6) |
| Unknown | 9163 | 0 (0–1) | 2 (1–2) | 0 (0–2) | 0 (0–0) | 1 (0–1) | 0 (0–1) | 5 (3–7) |
| **Primary Language** | | | | | | | | |
| English | 57879 | 0 (0–1) | 2 (1–2) | 0 (0–1) | 0 (0–0) | 1 (0–1) | 0 (0–1) | 4 (2–6) |
| Non-English | 5789 | 0 (0–1) | 2 (1–2) | 0 (0–0) | 0 (0–0) | 1 (0–1) | 0 (0–1) | 4 (2–6) |
| Unknown | 347 | 0 (0–1) | 2 (1–3) | 0 (0–2) | 0 (0–0) | 1 (0–2) | 0 (0–1) | 5 (4–8) |
| **Insurance Status** | | | | | | | | |
| Private | 18073 | 0 (0–1) | 1 (0–2) | 0 (0–1) | 0 (0–0) | 1 (0–1) | 0 (0–1) | 4 (2–6) |
| Medicare | 33789 | 0 (0–1) | 2 (1–2) | 0 (0–1) | 0 (0–0) | 1 (0–2) | 0 (0–1) | 4 (3–7) |
| Medicaid | 9149 | 0 (0–1) | 1 (0–2) | 0 (0–1) | 0 (0–0) | 1 (0–1) | 0 (0–1) | 4 (2–6) |
| Other | 3004 | 0 (0–1) | 1 (0–2) | 0 (0–1) | 0 (0–0) | 1 (0–1) | 0 (0–1) | 4 (2–6) |

**eFigure 2:** Calibration of SOFA-2 for ICU Mortality by Age Group

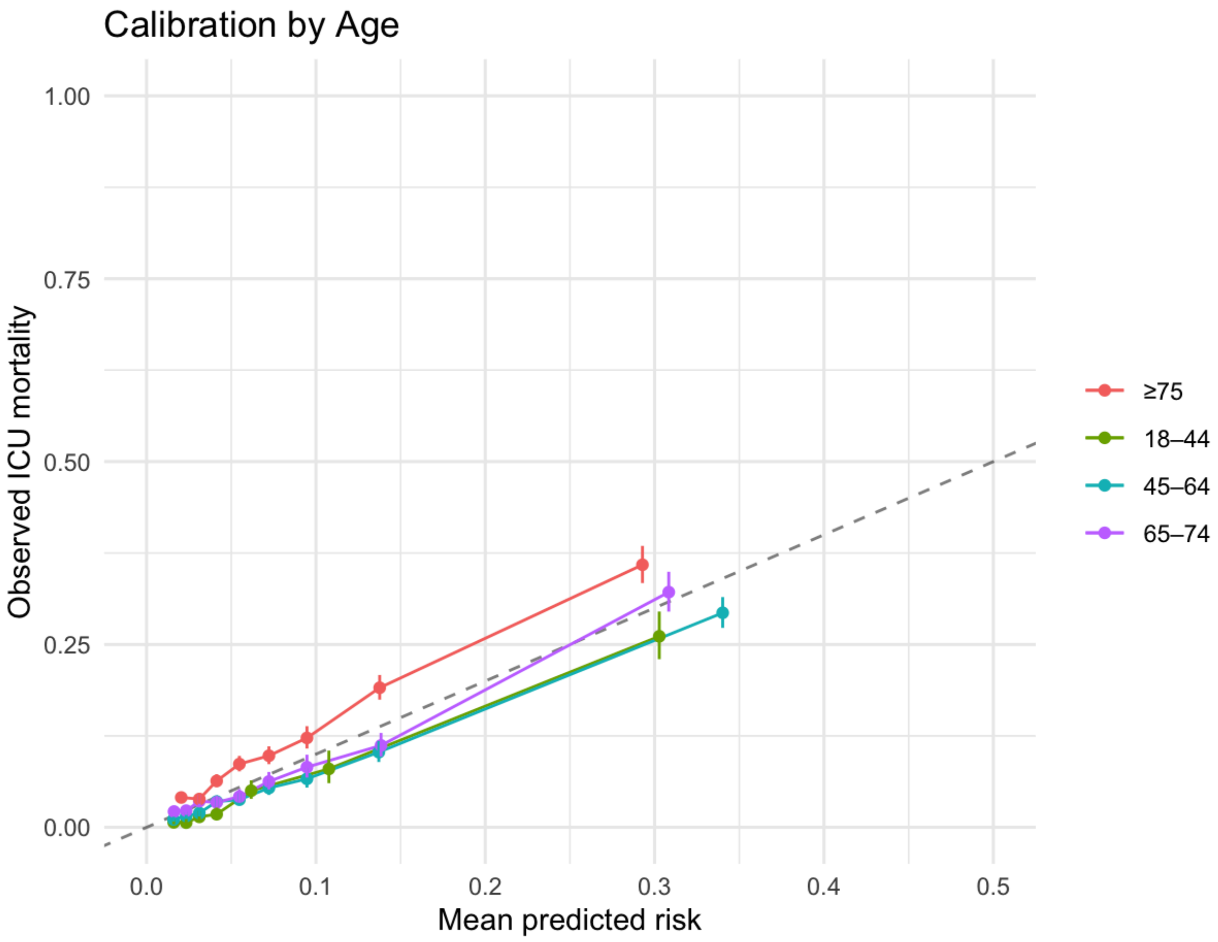

**eFigure 3:** Calibration of SOFA-2 for ICU Mortality by Sex

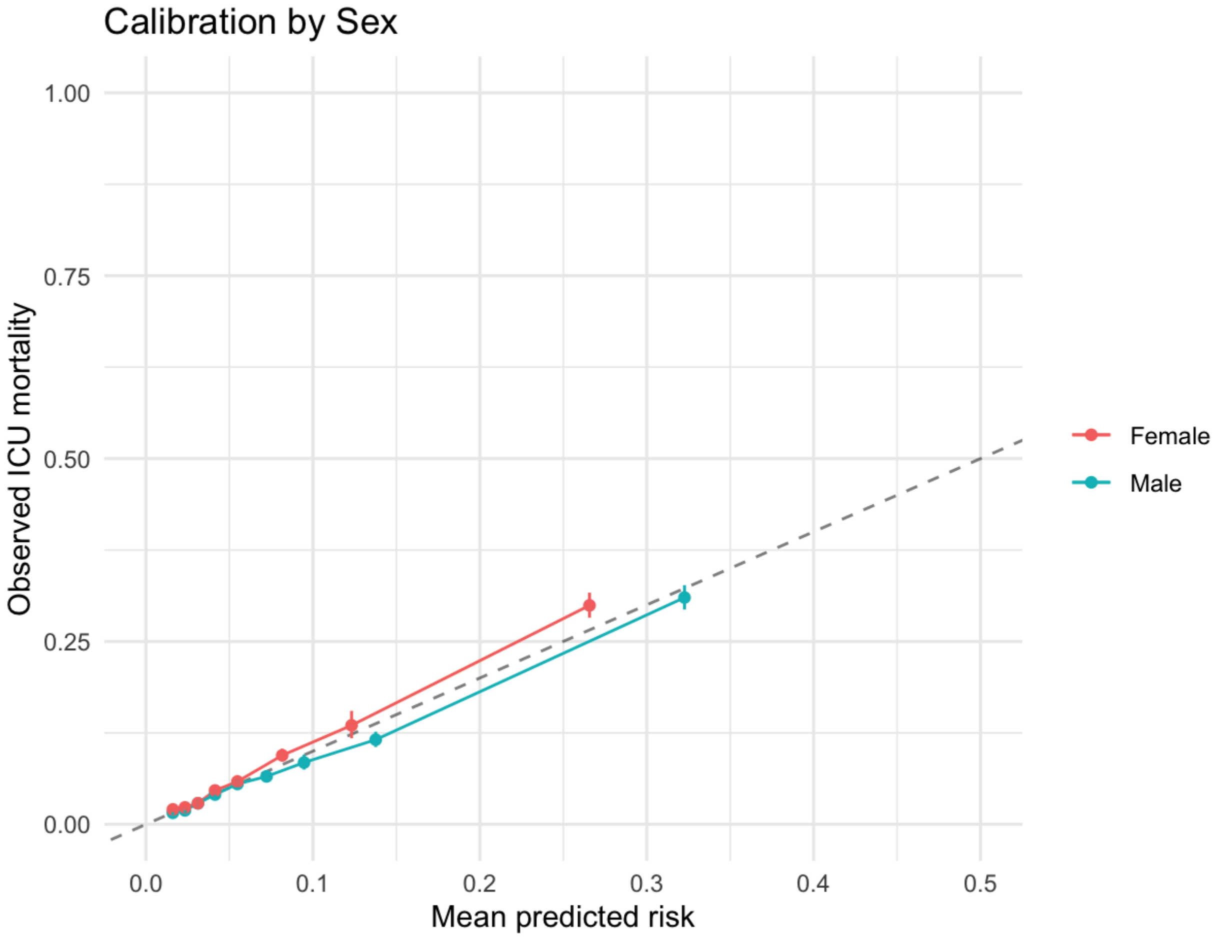

**eFigure 4:** Calibration of SOFA-2 for ICU Mortality by Race and Ethnicity

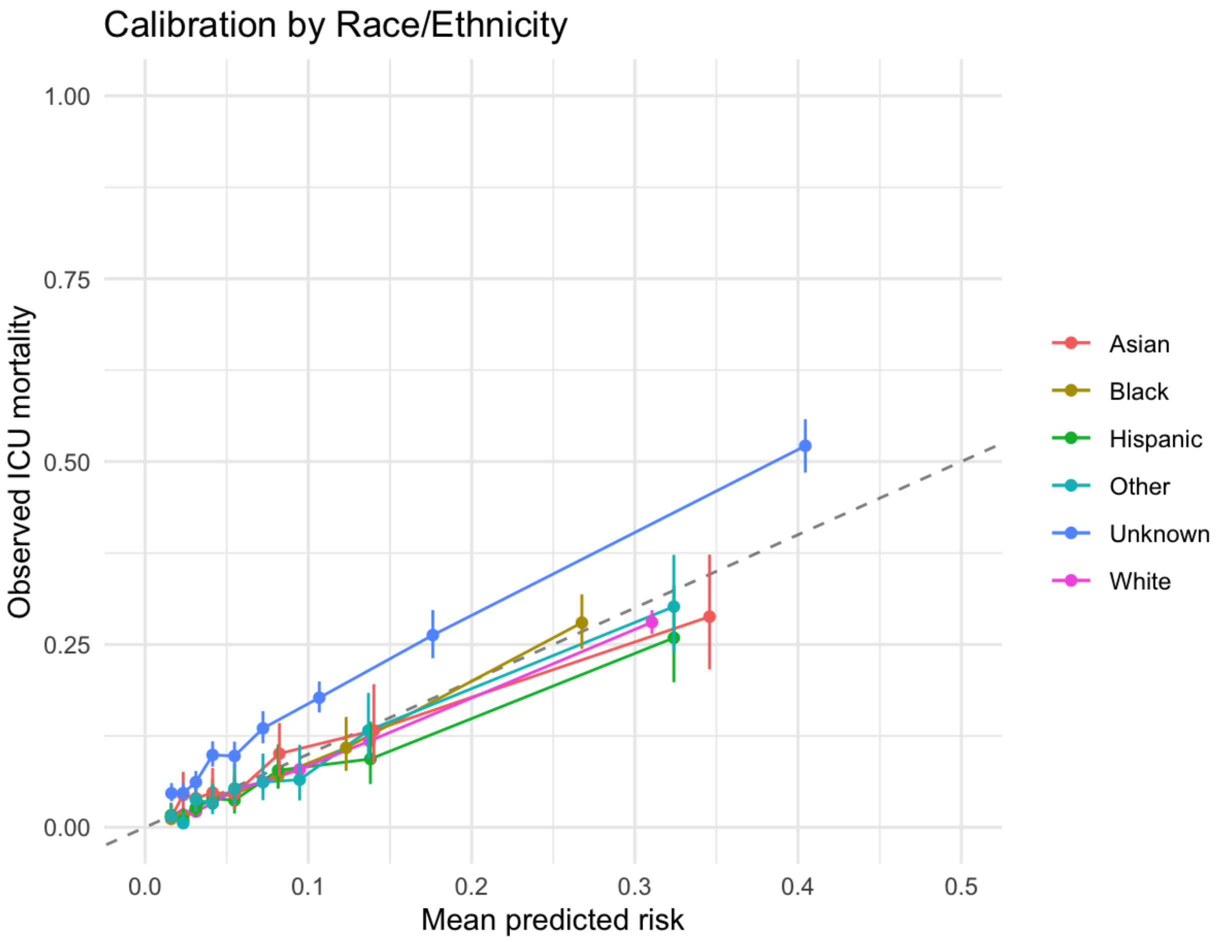

**eFigure 5:** Calibration of SOFA-2 for ICU Mortality by Primary Language

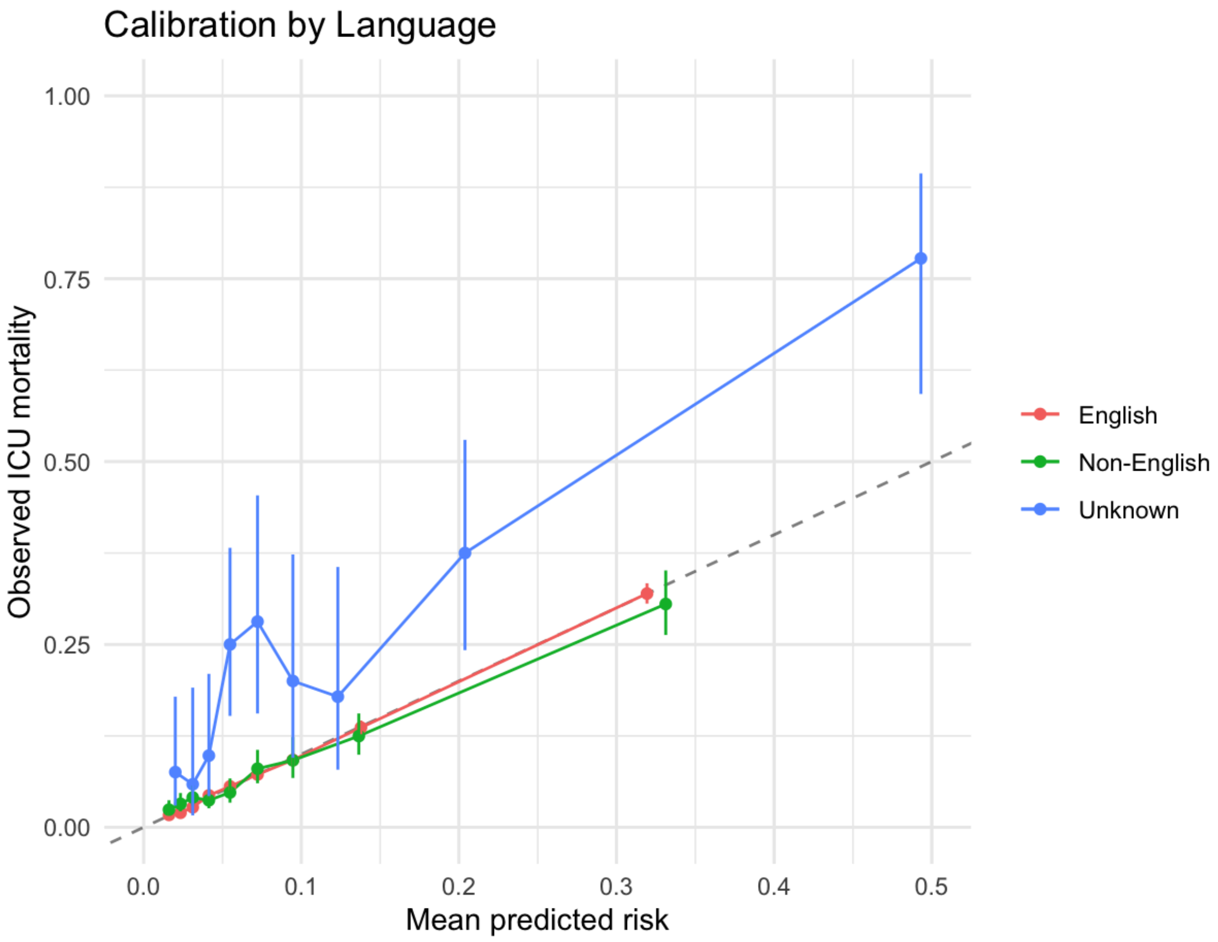

**eFigure 6:** Calibration of SOFA-2 for ICU Mortality by Insurance Status

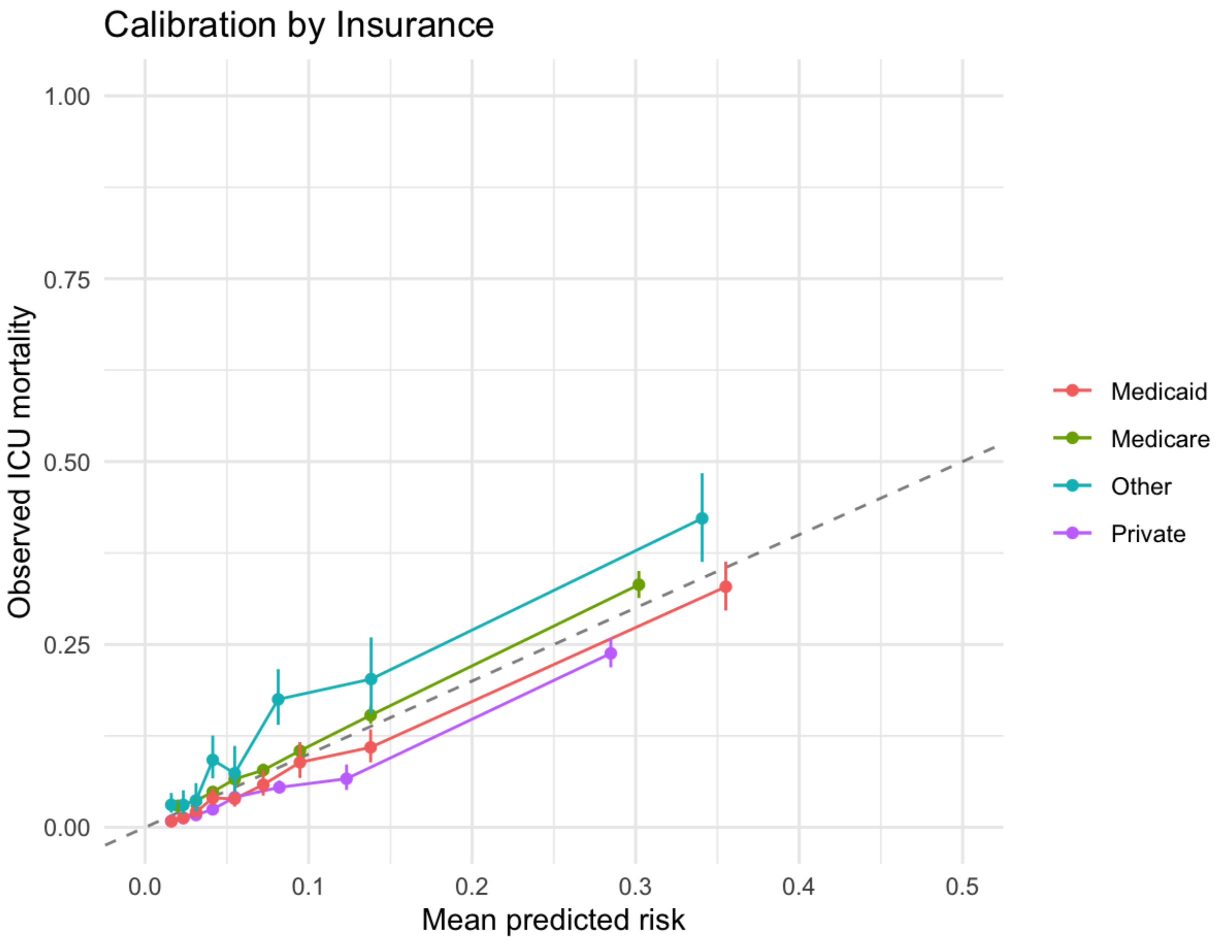

**eFigure 7:** Observed ICU Mortality by SOFA-2 Score Stratified by Race/Ethnicity

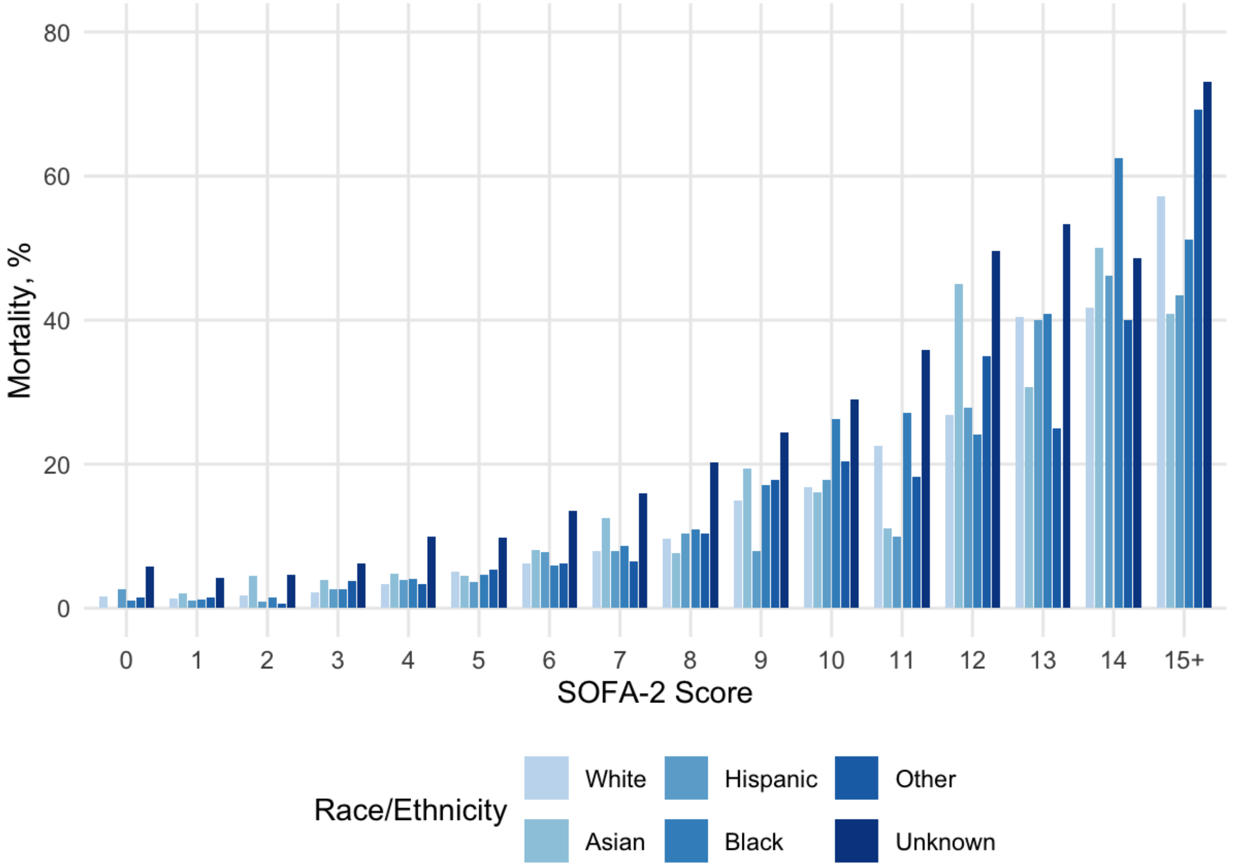

**eFigure 8:** Observed ICU Mortality by SOFA-2 Score Stratified by Primary Language

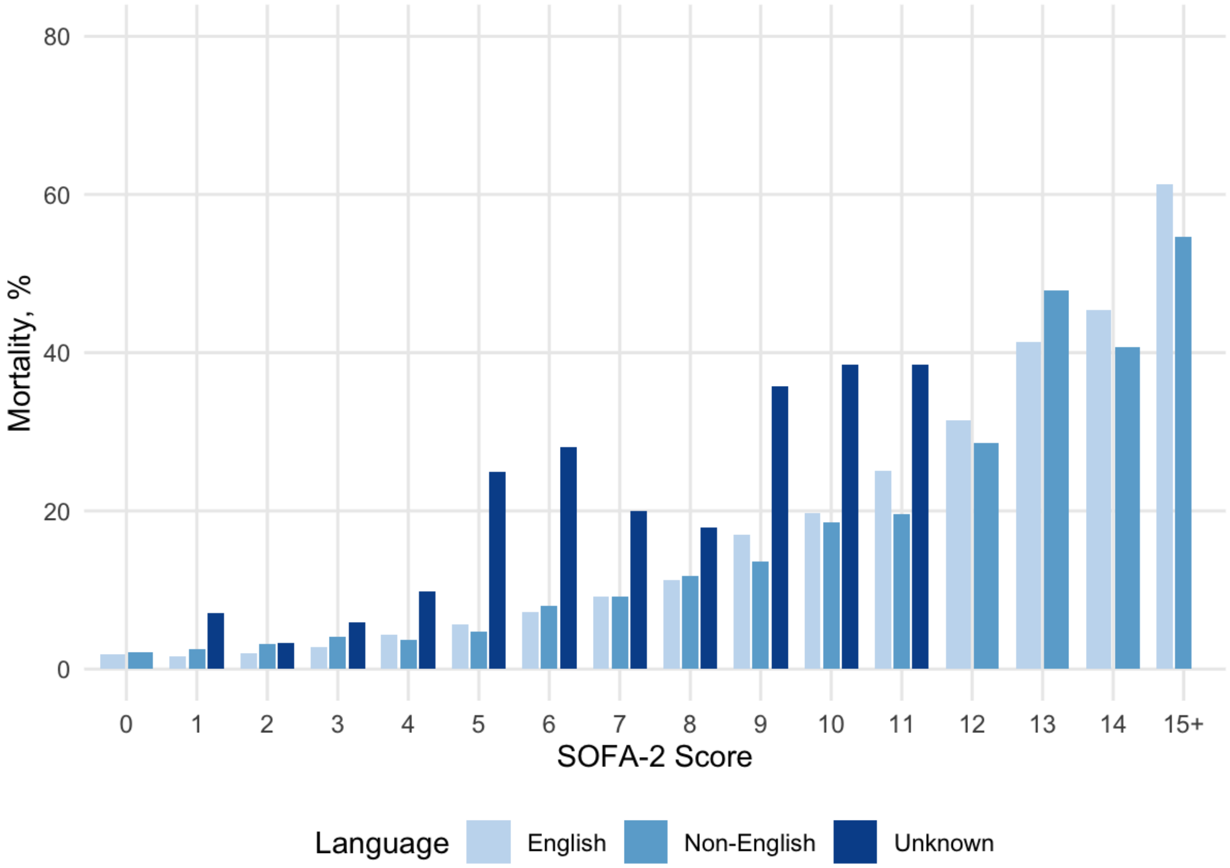

**eFigure 9:** Observed ICU Mortality by SOFA-2 Score Stratified by Insurance

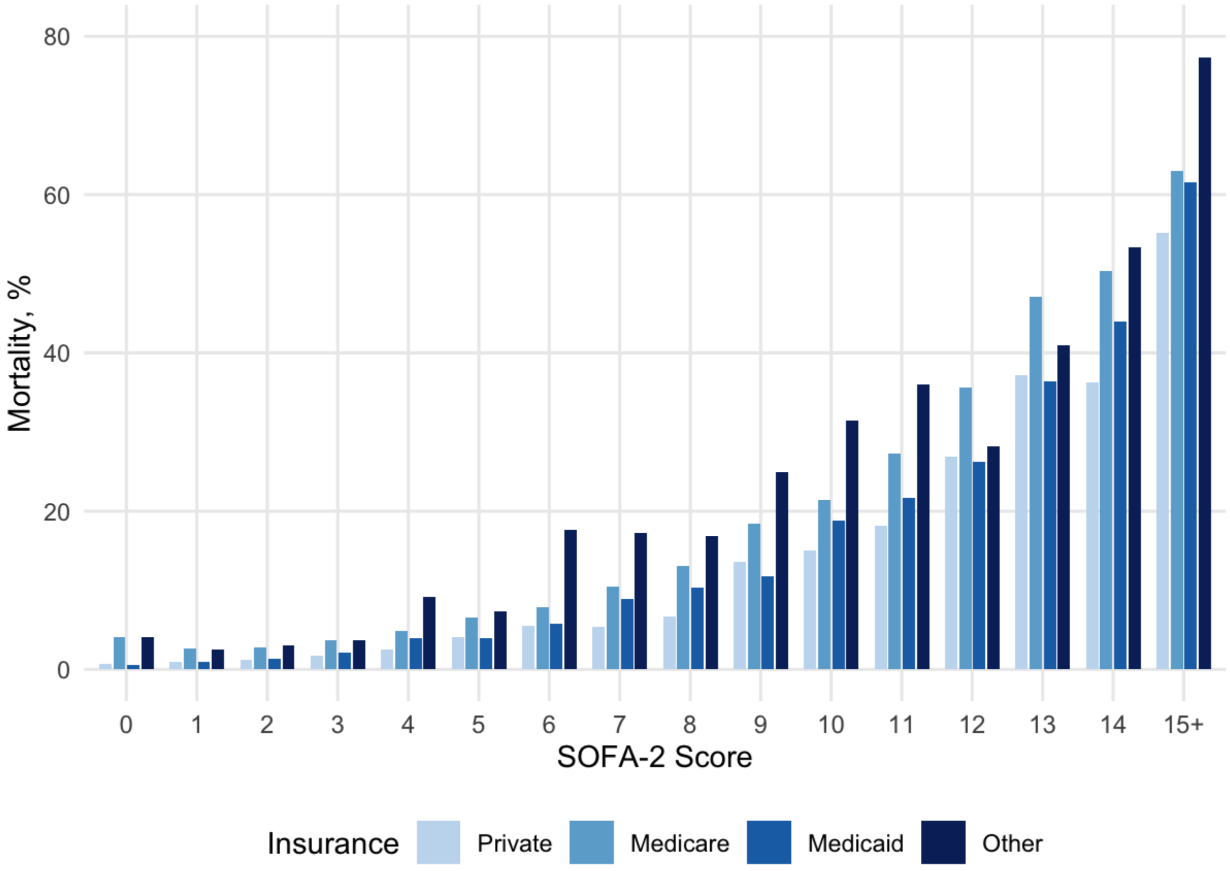
